# Geometric changes to the central nervous system organs at risk during chemoradiotherapy for locally advanced nasopharyngeal carcinoma

**DOI:** 10.1101/2022.07.01.22277168

**Authors:** Jing Li, Zeying Jiang, Wenyong Tan

**Author notes:** Author Contributions: Conceptualization, W.T., Z.J. and J.L.; methodology, W.T., Z.J. and J.L.; formal analysis, W.T., Z.J. and J.L.; writing—original draft preparation, J.L.; writing—review and editing, W.T.; funding acquisition, W.T. All authors have read and agreed to the published version of the manuscript. Funding: This research was funded by National Natural Science Funding of China (81974462). Institutional Review Board Statement: The study was conducted in accordance with the Declaration of Helsinki, and approved by the Institutional Review Board for the Hubei Cancer Hospital (No. 2010V011, Aug. 19, 2010). Informed Consent Statement: Informed consent was obtained from all subjects involved in the study. Data Availability Statement: The datasets used and/or analyzed during the current study are available from the corresponding author on reasonable request. (J.L.), (Z.J.). (W.T.).

## Abstract

**Background:** Considerable geometric changes to the organs at risk (OARs) have been reported during treatment with chemotherapy and intensity modulated radiotherapy (chemo-IMRT) for locally advanced nasopharyngeal carcinoma (LA-NPC). This study aimed to quantify geometric changes to the central nervous system-related OARs (CNS-OARs), during chemo-IMRT for LA-NPC.

**Methods:** This prospective study included 20 patients with LA-NPC, treated with chemo-IMRT. Patients underwent a planning computed tomography (CT-plan) scan with intravenous contrast, followed by six weekly scans without contrast (CT-rep). For CNS-OARs, including the spinal cord, brainstem, optic chiasm and nerves, the globes of the eye, lens, and inner ears, their volume loss, the center of mass (COM) displacement, and spatial deformation was compared among weeks, respectively. The correlation between organ volume reduction and patients’ weight loss was estimated.

**Results:** The volume of the brainstem, spinal cord, and the globe of left-and right-side eye averagely decreased by 2.6±2.3% (95% CI: 2.1%, 3.1%), 6.5±4.8% (5.6%,7.4%), 9.4±6.9% (8.1%, 10.6%) and 9.6±7.8% (8.2, 11.1%) respectively. The volume reduction of the spinal cord and that of the brainstem were significantly correlated with patients’ weight loss. For all OARs, the COM displacement was within 3 mm, except for the lower level of the spinal cord. The DSC value of the spinal cord, brainstem, and the globes of the eye was of >0.85 throughout treatment.

**Conclusions:** The volume and shape changes to the CNS-OARs during chemo-IMRT for NPC were quantifiable, which could be useful to refine radiation treatment protocols.

## 1. Introduction

Intensity modulated radiotherapy (IMRT) for nasopharyngeal carcinoma (NPC) is associated with better locoregional control and lower risk of toxicities than those associated with 2-dimensional radiotherapy (RT) [1]. However, radiotherapy planning and delivery are susceptible to errors, including those resulting from setup uncertainty and organ motion [2]. Imaging-guided RT (IGRT) can help minimize the impact of these errors. Radiotherapy assumes that the geometry of tumor and organs at risk (OARs) remains stable throughout treatment [3, 4]. However, previous studies have shown that the target volume (TV) and OAR geometry may undergo substantial changes during IMRT [5-12]. For example, previous studies of patients with head and neck cancer have revealed that a considerable TV shrinkage occurred over the course of treatment [5, 7-9]. In fact, for OARs such as the parotid gland, the volume decreased with consecutive radiation fractions [5,6], while its geometric center shifted medially by a few millimeters during RT [10]. Considering the priority of the central nervous system in the radiotherapy evaluation plan, it is necessary to evaluate whether it needs adaptive radiation to minimize the unnecessary irradiation during the treatment process [13]. Radiation-induced toxicities of the CNS-related OARs (CNS-OARs) can result in brain or spinal cord necrosis, blindness, and hearing loss, among others [14-17]. Previous studies have reported on the dose-volume effect in this context, without accounting for the geometric changes to the CNS-OARs [18]. However, during the definitive chemo-IMRT, the CNS-OARs such as the brainstem, spinal cord, and the globes of the eye may undergo geometric changes that result in dose uncertainty. This study aimed to quantify the geometric (volume, displacement, and shape) changes to the CNS-OARs, including several sub-regions of the cervical spinal cord to estimate the extended margin required for each CNS-OAR in IMRT planning, and to clarify the impact of these changes on the adaptive RT (ART) strategies for patients with locally advanced NPC (LA-NPC).

## 2. Materials and Methods

The whole study obeys the statement of Declaration of Helsinki, with the approval of the Institutional Review Board for the Hubei Cancer Hospital (No. 2010V011), and the need for informed consent was waived. Totally, twenty patients underwent concomitant chemo-IMRT, and among them seven patients received induction chemotherapy. During the posture fixation process, each patient performed the appropriate standard headrest model. Intravenous contrast planning computed tomography (CT-plan) scan and six repeat CT scans without contrast performed every five fractions (CT-rep) were acquired for each patient. Each CT-rep image was rigid registered with a bone match to its respective CT-plan, using purpose-created software, details of which have been previously described elsewhere [12]. Weekly data on patients’ weight were extracted from medical records.

### Delineations of CNS-OARs and spinal cord sub-regions

Seven CNS-OARs were delineated on each CT scan, specifically, the brainstem, spinal cord, globes of the eye, optic chiasm and nerves, lens, and inner ears, following international consensus guidelines [19], and the inner ear was defined as the cochlea [20]; Meanwhile, the cranial border of brain stem was identified as the bottom of lateral ventricle. The lower boundary of chiasm was recognized as the upper edge of pituitary gland and the upper bound is the uppermost edge of the optic chiasm visible in the image. The upper boundary of the spinal cord is defined as the upper edge of the odontoid process of the second cervical vertebra, and the lower boundary is 2cm from the lower edge of the clavicle head. The CT window width and level for contouring CNS-OARs were 300/40HU for soft structure and 1600/400HU for bone structure. Along the axial section, the spinal cord was divided into seven sub-regions based on bone marks; the anatomical margins of the cervical node levels were contoured on the CT images [21]. The sub-regions included the caudal (C1), cranial (C2S), and caudal (C2I) edge of the second cervical vertebra, and the caudal edge of the hyoid bone (Chb), cricoid cartilage (Ccc), cervical transverse vessels (Ctv), and suprasternal notch (Cns).

This division allowed to differentiate between treatment-related changes to the spinal cord and those to the lymph nodes. All delineations were performed by the same oncologist (WT) to minimize the effect of inter-observer differences.

### Geometric metrics

Geometric metrics included volume changes, positional shift, and shape variation estimates. The volume parameter and center of mass (COM) of each CNS-OAR were calculated using the WALDMATC software [12]. The positional shift was quantified with the mean displacement (M), the systematic (Σ) and random (δ) error in the left-right (LR), cranial-caudal (CC), and anterior-posterior (AP) directions, respectively [22]. The displacement of COM in each CNS-OAR was calculated in the LR, CC, and AP direction, and the 3-dimensional displacement vector (3D-vector displacement) was calculated using the previously reported formula as 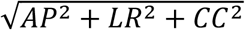. Meanwhile, since the position of the cervical spinal cord in the axial section of each anatomical part is determined according to the bony imaging landmarks, there is no CC displacement for spinal cord, and the 2-dimensional displacement vector of the spinal cord was estimated with the formula of 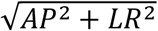 [23]. The margins of planning organ at risk volume (PRV) were estimated along the three axes, using the formula, 1.3Σ+0.5δ [24]. When compared the same OAR in CT_plan with CT__repeat_, the volume in CT__plan_ (V__plan_) and CT__rep_ (V__repeat_) as their union volume (V__union_) and intersection volume [25] and (V_inter) were calculated. Shape-related parameters included the Dice similarity index coefficient (DSC) and overlapping index (OI) were estimated as the formula of 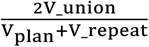 [25] and 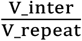 [26] respectively. The shortest perpendicular distance (SPD) and its standard deviation (SD) were also automatically calculated, using the method previously reported by Heimann et al [27]. The same parameters were used to evaluate the changes to the sub-regions of the cervical spinal cord.

### Statistical analysis

The Shapiro-Wilk and Kolmogorov-Smirnov tests were used to test the assumptions of the normality of distribution of the data, respectively. The volume loss of each CNS-OARs was compared among treatment weeks, with the first week volume loss as a reference. According to whether the data followed the assumptions of normality distribution, repeated measures analysis of variance (RM-ANOVA) or non-parametric Friedman’s test was applied to test whether a difference occurred between weeks in each organ. The two-tailed paired Student’s t test or Wilcoxon test was used to assess the significance between each paired comparison, and a Bonferroni method was used to correct the above paired test. The rates of volume reduction and weight loss of CNS-OARs were estimated by linear regression analysis. A p-value of <0.05 was considered indicative of a statistically significant finding. The Statistical Package for the Social Sciences (SPSS), version 20.0 (IBM Corporation, Armonk, NY, USA) was used for all statistical analyses.

## 3. Results

### 3.1. Volume changes

Following IMRT, the volume of the spinal cord and brainstem decreased by 6.5±4.8% and 2.6±2.3%, respectively (Table 1). The rate of volume reduction of these two structures decreased over time. However, volume changes to the optic nerve and chiasm, lens and the globes of eyes, and inner ears followed a different pattern. For the spinal cord, brainstem, and the globes of the eye, volume reduction after the first five IMRT fractions was 2.8±3.1%, 1.1±1.5%, and 4.6±6.5%, respectively, while the corresponding values after 30 IMRT fractions were 9.6±5.2%, 3.8±3.2%, and 13.1±6.3% (Table 1), respectively. Among these organs, the globes of the eye, and the spinal cord and brainstem first presented with distinct volume reduction at 2th and 3rd week, when compared with the first week, respectively. (p=0.001, p=0.008, p=0.007, respectively). Volume reduction of the brainstem (R2=0.106, p=0.000) and that of the spinal cord (R2=0.378, p=0.000) were correlated with patients’ weekly weight loss (Figure 1A, 1B). In contrast, the volume reduction of the globes of the eye did not correlate with patients’ weight loss (Figure 1C). Since the optic nerve volume change data was not meet the normal distribution, we did not perform linear regression calculations between the optic nerve volume loss and patients’ weekly weight loss.

**Table 1.** volume changes of CNS-related OARs among different weeks during chemoIMRT for NPC

| OARs | W1 | W2 | W3 | W4 | W5 | W6 | Mean |
| --- | --- | --- | --- | --- | --- | --- | --- |
| Brainstem | -1.1±1.5<br>(-1.7, -0.4) | -1.7±1.5<br>(-2.5, -1.0) | -2.4±1.7<br>(-3.2, -1.6) | -2.8±2.1<br>(-3.8,-1.8) | -3.5±2.4<br>(-4.6, -2.4) | -3.8±3.2<br>(-5.3, -2.3) | -2.6±2.3<br>(-3.1, 2.1) |
| Spinal cord | -2.8±3.1<br>(-4.2, -1.3) | -4.4±3.5<br>(-6.1, -2.8) | -6.0±4.1<br>(-7.9, -4.1) | -7.6±4.8<br>(-9.8, -5.3) | -9.6±5.2<br>(-12.0, -7.2) | -9.6±5.2<br>(-12.0, -7.2) | -6.5±4.8<br>(-7.4, -5.6) |
| Optic Chiasm | -5.6±10.6<br>(-10.5, -0.6) | -6.4±19.4<br>(-15.4, 2.7) | -9.9±19.8<br>(-19.1, --.6) | -11.0±20.1<br>(-20.4, -1.6) | -14.3±21.9<br>(-24.6, -4.1) | -15.4±21.6<br>(-25.5, -5.3) | -10.4±19.2<br>(-13.9, -6.9) |
| Optic nerves | 1.2±22.3<br>(-6.0, 8.4) | -1.2±22.6<br>(-6.0, -8.5) | -1.0±26.1<br>(-9.3, 7.4) | -1.0±26.9<br>(-7.7, 9.6) | -1.0±27.7<br>(-9.8, 7.9) | -1.6±22.8<br>(-8.9, 5.7) | -0.0±24.6<br>(-3.2, -3.1) |
| Eye globes | -4.6±6.5<br>(-6.7, -2.5) | -7.2±6.8<br>(-9.3, -5.0) | -9.2±6.8<br>(-11.4, -7.0) | -10.3±7.5<br>(-12.6, -7.9) | -12.6±6.8<br>(-14.8, -10.5) | -13.1±6.3<br>(-15.1, -11.1) | -9.5±7.4<br>(-10.4, -8.6) |
| Lens | 2.6±30.0<br>(-7.0, 12.1) | -0.2±38.5<br>(-12.5, -12.1) | 1.1±53.5<br>(-16.0, -18.2) | -0.2±51.7<br>(-16.7, -16.4) | -1.6±50.3<br>(-14.4, 17.7) | -0.9±49.6<br>(-16.8, -15.0) | -0.7±45.9<br>(-5.2, 6.5) |
| Ear inner | -10.1±21.7<br>(-17.0, -3.1) | -10.8±25.3<br>(-18.9, -2.7) | -10.2±23.1<br>(-17.6, -2.8) | -11.0±24.4<br>(-18.8, -3.1) | -11.0±27.0<br>(-19.7, -2.4) | -5.1±38.2<br>(-17.3, 7.1) | -9.7±27.0<br>(-13.1, -6.3) |

Abbreviations: CNS, central nervous system; OARs, organ at risks; chemo-IMRT, chemotherapy combined with intensity-modulated radiotherapy; NPC, nasopharyngeal carcinoma; SD, standard deviation; CI, confidence interval.

**Figure 1.**
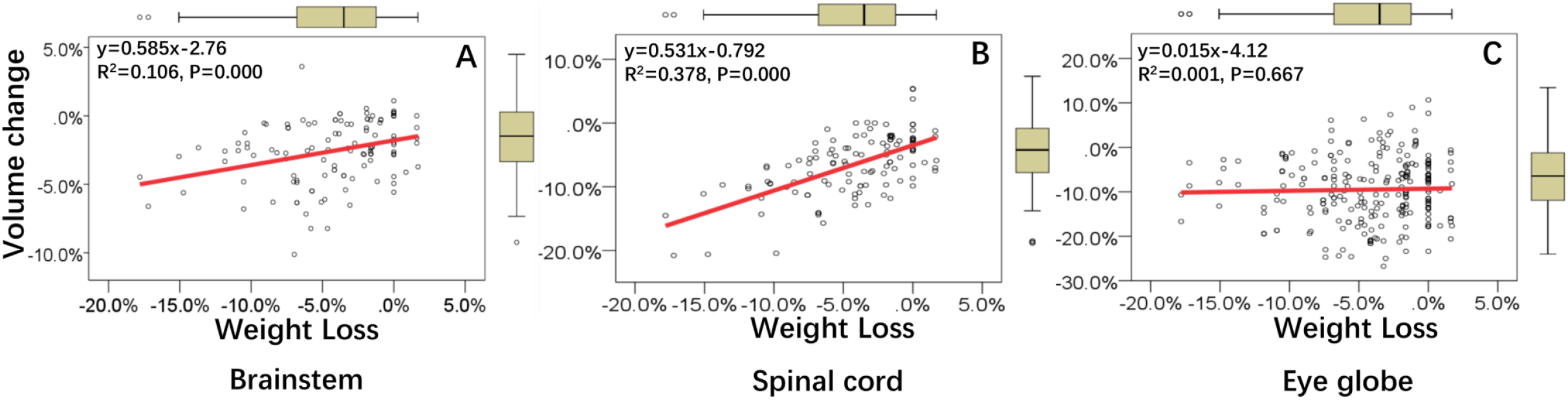
Association between weight loss and volume changes to the (A) brainstem, (B) spinal cord, and (C) eye globe, derived from linear regression. Regression equations are presented in the upper left corner of each panel. The lines show the means, and the error bars show the 95% confidence intervals.

### 3.2. Positional displacement

The spinal cord was displaced in the AP and CC directions by 0.30 mm and 0.02 mm, respectively (Table 2). The brainstem recorded displacement in all directions within the range of 0.11-0.13 mm. In the visual system, the optic chiasm shifted in the LR direction by 0.15 mm; the optic nerves shifted toward both the left and right side in the AP direction by 0.27 mm and 0.33 mm, respectively. However, the globes of the eye and the lens presented heterogeneous patterns of displacement. For example, the maximum displacement of the right lens in the CC direction was larger than that of the left lens in the AP direction. Similarly, the globes of the left and right eye were displaced in the CC direction by 0.25 mm and 0.11 mm, as well as those of 0.09mm and 0.18mm in AP direction, respectively. Concurrently, both inner ears tended to shift in the LR direction within 0.2 mm. The median 3D-vector displacement of all CNS-OARs was within 3 mm (Figure 2A).

**Table 2.**
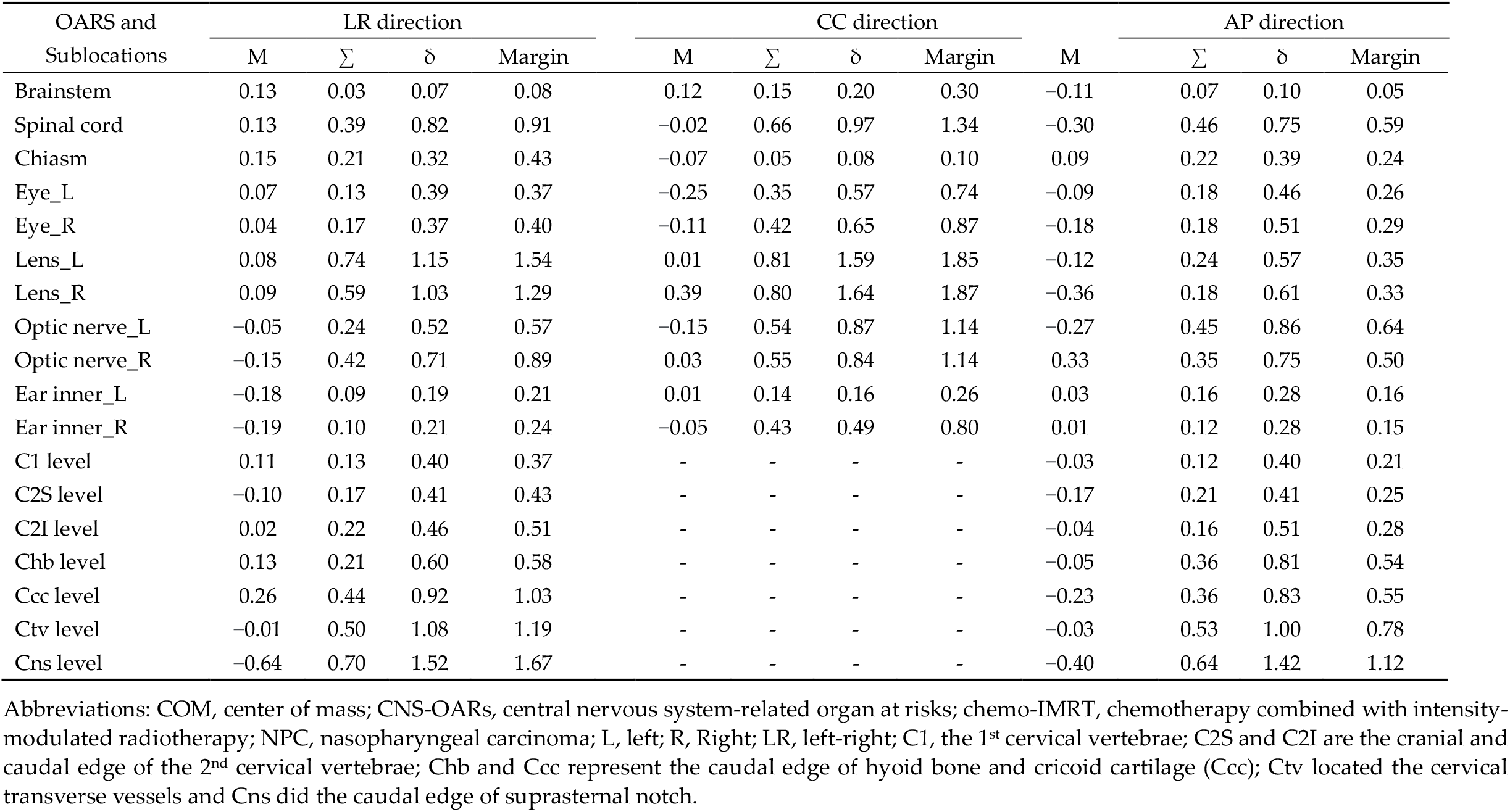
Mean, system, and random errors and the estimated margin of the COM to the CNS-OARs (mm)

**Figure 2.**
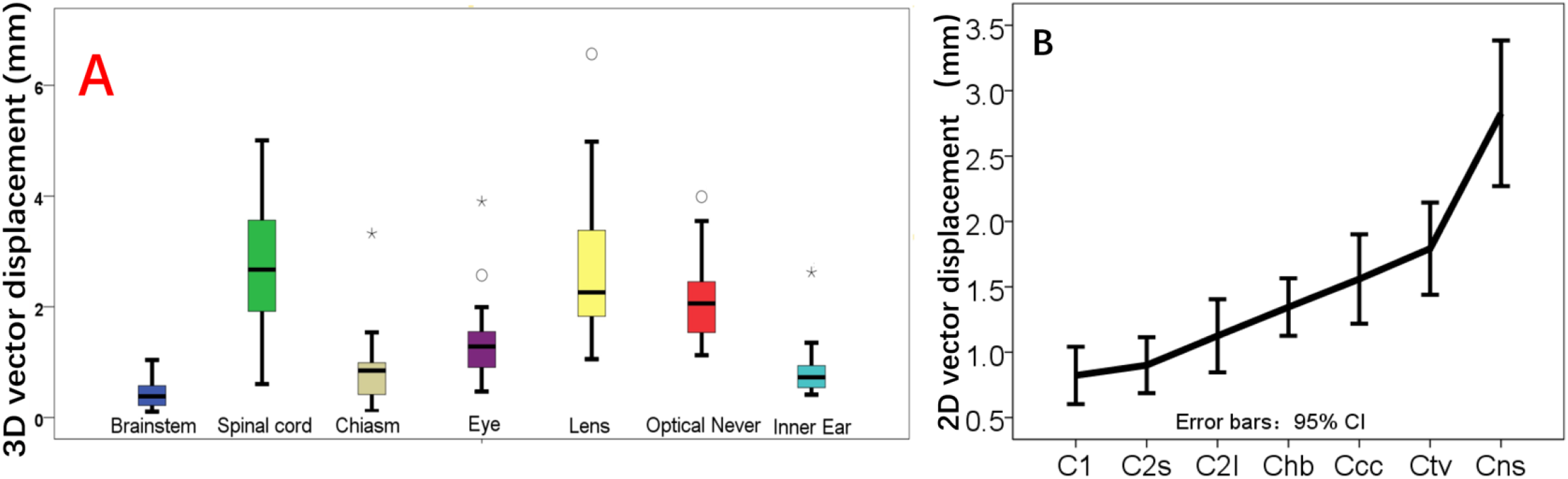
The box-and-whisker plot of three-dimensional vectors (3D-vector) of the center of mass (COM) displacement for the central nervous system-related organs at risk (CNS-OARs). The line in the box indicates the mean value and the error bars show the 95% confidence intervals (A). The two-dimensional vectors (2D-vector) of the COM displacement in the spinal cord subregions (B). The lines show the means, and the error bars show the 95% confidence intervals.

Although the 2D-vector displacement of the spinal cord sub-regions was of <3 mm (Figure 2B), the spinal cord sub-regions showed heterogeneous patterns of displacement (Table 2). Specifically, the upper spinal cord (at the C1, C2, C2l, and Chb levels), corresponding to the level II of the neck nodal regions [21], presented displacement in the range of 0.02-0.13 mm and 0.03-0.17 mm in the LR and AP directions, respectively. While the lower spinal cord regions (at the Ccc, Ctv, and Cns levels), corresponding to the level III and IV of the neck lymph nodes, presented displacement in the range of 0.01-0.64 mm and 0.03-0.40 mm in the LR and AP directions, respectively. The value of the 3D-vector displacement of the lower regions of the spinal cord was greater than that of the upper regions. In addition, the upper 95% confidence interval of the lower regions of the spinal cord was above 3mm. For all CNS-OARs, Σ of displacement values were of <1 mm and the corresponding δ values were of <1.7 mm in all directions (Table 2).

### 3.3. Shape changes

Both the OI and DSC values for the spinal cord, brainstem, and the globes of the eye were of 0.7 throughout treatment; the corresponding values for the lens and optic nerves were <0.7, suggesting considerable geometric variation (Table 3). The mean SPD of CNS-OARs was of <1 mm (0.46-0.89 mm) during treatment, except for the globes of the eye (0.83-1.01 mm), lens (0.83-1.01 mm), and optic nerves (0.99-1.38 mm). The SD of SPD was in the range of 0.54-0.62 mm and 0.85-1.03 mm for the inner ears and optic chiasm, respectively. The corresponding values for the brainstem, spinal cord, the globes of the eye, optic nerve, and lens were 0.89-1.18 mm, 0.98-1.24 mm, 1.07-1.24 mm, 1.22-1.62 mm, and 1.23-1.85 mm, respectively.

**Table3.** The DSC and SPD of CNS-OAR during chemo-IMRT to NPC patients (mean ±SD, 95% CI)

| OARs | W1 | W2 | W3 | W4 | W5 | W6 | Mean |
| --- | --- | --- | --- | --- | --- | --- | --- |
| DSC |  |  |  |  |  |  |  |
| Brainstem | 0.95±0.02<br>(0.94, 0.96) | 0.95±0.02<br>(0.94, 0.95) | 0.94±0.02<br>(0.94, 0.95) | 0.94±0.03<br>(0.93, 0.95) | 0.94±0.02<br>(0.93, 0.94) | 0.93±0.02<br>(0.92, 0.94) | 0.94±0.02<br>(0.93, 0.94) |
| Spinal cord | 0.87±0.05<br>(0.84, 0.89) | 0.87±0.08<br>(0.83, 0.90) | 0.87±0.04<br>(0.86, 0.89) | 0.85±0.06<br>(0.82, 0.88) | 0.85±0.06<br>(0.83, 0.88) | 0.87±0.03<br>(0.86, 0.88) | 0.86±0.06<br>(0.85, 0.87) |
| Optic chiasm | 0.80±0.12<br>(0.75, 0.86) | 0.77±0.15<br>(0.70, 0.84) | 0.78±0.13<br>(0.72, 0.84) | 0.78±0.13<br>(0.72, 0.84) | 0.73±0.12<br>(0.67, 0.78) | 0.72±0.12<br>(0.67, 0.78) | 0.76±0.13<br>(0.74, 0.79) |
| Optic nerves | 0.80±0.12<br>(0.75, 0.86) | 0.77±0.15<br>(0.70, 0.84) | 0.78±0.13<br>(0.72, 0.84) | 0.78±0.13<br>(0.72, 0.84) | 0.73±0.12<br>(0.67, 0.78) | 0.72±0.12<br>(0.67, 0.78) | 0.76±0.13<br>(0.74, 0.79) |
| Eye globes | 0.91±0.04<br>(0.90, 0.92) | 0.90±0.03<br>(0.89, 0.91) | 0.89±0.05<br>(0.87, 0.90) | 0.88±0.05<br>(0.86, 0.89) | 0.88±0.05<br>(0.86, 0.89) | 0.87±0.05<br>(0.86, 0.89) | 0.89±0.05<br>(0.88, 0.89) |
| Lens | 0.61±0.24<br>(0.53, 0.68) | 0.58±0.20<br>(0.51, 0.64) | 0.50±0.23<br>(0.42, 0.57) | 0.51±0.19<br>(0.45, 0.58) | 0.47±0.20<br>(0.41, 0.54) | 0.52±0.18<br>(0.46, 0.58) | 0.53±0.21<br>(0.50, 0.56) |
| Ear inner | 0.81±0.12 | 0.77±0.11 | 0.78±0.11 | 0.75±0.11 | 0.76±0.09 | 0.76±0.10 | 0.77±0.11 |

|  | (0.77, 0.85) | (0.73, 0.81) | (0.75, 0.82) | (0.71, 0.78) | (0.73, 0.79) | (0.73, 0.80) | (0.76, 0.79) |
| --- | --- | --- | --- | --- | --- | --- | --- |
| SPD (mm) |  |  |  |  |  |  |  |
| Brainstem | 0.58±0.27 | 0.65±0.20 | 0.66±0.20 | 0.72±0.30 | 0.74±0.21 | 0.80±0.21 | 0.69±0.24 |
|  | (0.46, 0.71) | (0.55, 0.74) | (0.56, 0.75) | (0.58, 0.86) | (0.64, 0.84) | (0.70, 0.90) | (0.65, 0.73) |
| Spinal cord | 0.76±0.31 | 0.79±0.53 | 0.74±0.21 | 0.89±0.39 | 0.84±0.35 | 0.72±0.17 | 0.79±0.35 |
|  | (0.61, 0.90) | (0.54, 1.03) | (0.64, 0.83) | (0.71, 1.08) | (0.67, 1.00) | (0.64, 0.80) | (0.73, 0.85) |
| Optic chiasm | 0.60±0.39 | 0.73±0.59 | 0.74±0.57 | 0.68±0.52 | 0.75±0.42 | 0.76±0.45 | 0.71±0.49 |
|  | (0.41, 0.78) | (0.46, 1.01) | (0.47, 1.01) | (0.43, 0.92) | (0.55, 0.95) | (0.55, 0.97) | (0.62, 0.80) |
| Optic nerves | 0.99±0.66 | 1.16±0.73 | 1.33±0.99 | 1.31±0.60 | 1.34±0.62 | 1.38±0.70 | 1.25±0.73 |
|  | (0.77, 1.21) | (0.92, 1.40) | (1.01, 1.64) | (1.12, 1.50) | (1.14, 1.53) | (1.15, 1.61) | (1.16, 1.35) |
| Eye globes | 0.83±0.48 | 0.83±0.27 | 0.94±0.41 | 1.00±0.47 | 0.98±0.44 | 1.01±0.47 | 0.93±0.43 |
|  | (0.68, 0.99) | (0.75, 0.92) | (0.81, 1.08) | (0.85, 1.15) | (0.84, 1.12) | (0.86, 1.15) | (0.88, 0.99) |
| Lens | 0.83±0.48 | 0.83±0.27 | 0.94±0.41 | 1.00±0.47 | 0.98±0.44 | 1.01±0.47 | 0.93±0.43 |
|  | (0.68, 0.99) | (0.75, 0.92) | (0.81, 1.08) | (0.85, 1.15) | (0.84, 1.12) | (0.86, 1.15) | (0.88, 0.99) |
| Ear inner | 0.46±0.30 | 0.57±0.33 | 0.54±0.24 | 0.61±0.28 | 0.56±0.28 | 0.57±0.35 | 0.55±0.30 |
|  | (0.36, 0.56) | (0.46, 0.68) | (0.46, 0.62) | (0.52, 0.70) | (0.47, 0.65) | (0.46, 0.69) | (0.51, 0.59) |

Abbreviations: DSC, Dice similarity coefficient; SPD, the shortest perpendicular distance; CNS-OARs, central nervous system-related organ at risks; chemo-IMRT, chemotherapy combined with intensity modulated radiotherapy; NPC, nasopharyngeal carcinoma; SD, standardized deviation; CI, confidence interval. Both the DSC and SPD could represent the shape uncertainties of CNS-OARs during the ChemoIMRT.

## 4. Discussion

In this study, we quantified the geometric changes to the CNS-OARs throughout the course of chemo-IMRT for LA-NPC. To the best of our knowledge, it is the first to demonstrate the volume of brainstem and spinal cord decreased over time during IMRT. And positional displacement of spinal cord at the lower neck was much larger than that in the upper neck. Though the volume and position displacement of CNS-OARs with small volume such as the lens, optical nerve, chiasm, and inner ear were rather unsignificant, the overlapping between the following repeat CT and initial planning CT was not adequate for the IMRT. These findings suggest that more precautions should be taken when the tumor invaded base of skull and especially abutted to these CNS-OARs. Adaptive replanning might be one of useful strategies to mitigate the risk of over radiation dose to CNS-OARs for head and neck cancer.

Previous studies have shown no significant changes to the volume of the spinal cord during chemo-IMRT [28]. However, our study has shown that such changes may occur, including to the spinal cord and brainstem during the third week of chemo-IMRT. And the changes to these two CNS-OARs followed a time trend as parotid gland did [7, 29, 30]. This phenomenon highlights the need to pursue ART during the third week of treatment. Meanwhile, although there are changes in the volume of the inner ear, as a small-volume organ, its volume change is greatly affected by the delineation error, rather than the volume change caused by treatment.

Alongside the uncertainty associated with image registration, changes to the organ volume may affect its placement. The values of the 3D-vector displacement, systematic, and random displacement errors of the COM of each OAR may help quantify their positional shift during chemo-IMRT. In the present study, the 3D-vector displacements of all OARs were within 3-5 mm, with the systematic and random error values of <3 mm. As the cervical spinal cord is a serial OAR with longitudinal length of >10 cm, single-point assessments fail to fully present the overall changes. A previous study has reported the maximum COM displacement of the spinal cord as within 3.5 mm and 5.6 mm at the C1 and C6 levels, respectively, with most shifts occurring in the LR direction [31]. Consistent with these findings, in our study, the upper cervical region of the spinal cord presented a smaller average COM displacement value in the axial section than did the lower cervical region (within 1.5 mm and 3 mm, respectively). Robar et al. reported the average COM displacement of the brainstem as 2.9 mm and 3.4 mm in the superior and inferior sections, respectively [31].

In the present study, the overall shift of the brainstem was within 2 mm. Relative to the 3D-vector displacement of brainstem and spinal cord, these of the optic nerves, chiasm, and inner ears was within 2 mm. However, due to the small volume of these organs, the displacement estimates may have resulted from poor imaging quality rather than their deformation. In addition, we evaluated the COM displacement on the effect of intra-fraction anatomical changes, although the intrafraction error and neck curvature will also affect the results to a certain extent. The cone beam computed tomography (CBCT) can also be used to roughly evaluate local COM displacement. The present study used the same setting parameters as the current treatment to collect repeated CT and similar rigid registration methods. The obtained repeated CT can monitor the anatomical changes that CBCT cannot observe, especially the volume and shape changes. Individualized foam pads could be used to reduce the dose uncertainty caused by changes in the neck curvature, as a retrospective study, we will further study the anatomical changes of the organs during the long course of treatment after more precise control of the neck curvature in the posture fixation process.

The DSC index values larger than 0.7 represent excellent overlap concordance [25]. In the present study, the shape of the spinal cord, brainstem, and the globes of the eye maintained the DSC value of >0.7 throughout IMRT. However, the DSC value of the CNS-OARs with small volume including the optical nerves, chiasm and lens was of <0.8 throughout treatment, likely due to the nuanced locations among different CT scanning, the delineation uncertainties, and the motion of the lens during intra-fractionation RT. The SPD and SD metrics were used to describe the surface changes of the OARs; these indices were previously used to assess spatial geometric changes to the TV. And the SPD and SD of TV as 4.6 mm and 3.7 mm during IMRT, respectively [12]. In the present study, the SPD and SD values of the CNS-OARs were within 2 mm. International guidelines recommend a 5-mm extended margin for OARs during the treatment of head and neck cancer [19]. However, some studies have proposed a 3-mm margin for PRV [32, 33]. From our results, the individualized PRV margin to each CNS-OARs might be more reasonable. Few studies have reported dosimetry changes to the CNS-OARs with a 3-mm extended margin. Liu et al. have validated the feasibility of a 3-mm margin for spinal cord and brainstem dosimetry. Meanwhile, a 5-mm margin was reported by Cheng et al. as required for radiation delivered to the lower region of the cervical spinal cord [8]. For small-volume OARs such as the optic nerves and chiasm, lens, and inner ears, the PRV margin should be sufficient to account for geometric uncertainties associated with rigid registration during imaging-guided IMRT. A dosimetry study revealed that during imaging-guided IMRT, the ipsilateral optic nerve received D1% of 7.5 Gy, with the cumulative maximum dose value of >60 Gy, given a 3-mm margin to bilateral optic nerves in NPC patients; this setup has been associated with high risk of radiation-induced optic neuropathy in the ipsilateral optic nerve [34]. ART may help clinicians manage the discrepancies between the planned and delivered radiation dose to the TV and OARs, as it may account for the potential geometric changes during RT [35]. The replanning strategy, which currently attracts limited consensus in clinical practice, could help accurately deliver the radiation dose to the TV while limiting the exposure of the OARs. For NPC patients, IMRT with replanning has been shown to improve the dosimetry of the parotoid glands and decrease the risk of toxicity, including that of xerostomia [36-38]. The loss of volume in the parotid gland tends to occur in the first 2-4 weeks of IMRT [6, 9], suggesting that ART in the early stages might benefit patients with head and neck cancer. A study has reported the both the upper and lower cervical region of the spinal cord experience lateral neck diameter and slice surface area value reduction at the end of RT course in patients with head and neck cancer [39]. However, in the study, no significant correlation observed between the two-dimension volume changes or weight loss, which defined as the difference value between the maximum dose that delivered and planned to spinal cord. Another study also showed three in thirty-one for head and neck patients need an adaptive replan radiotherapy to avoid excessive dose exposure for the D0.1cc to spinal cord [40]. Meanwhile, this study also reported an irrelevant relationship between spinal cord volume changes and the its dose increase. As reported in the present study, the brain stem and spinal cord showed volume decrease over time, and the volume change of the two organs correlated with patients’ weekly weight loss, suggesting that the third or fifth week, as well as weekly weight loss might be an indicator for triggering anatomic based ART, and whether an anatomical changing based ART will translate into dosemetic and/or clinical benefit warrant to the further study.

Though this study quantified the nuanced geometric changes for majority of CNS-OARs throughout the course of chemoIMRT for NPC, which may help refine routine IMRT and ART planning for LA-NPC. However, this study has some limitations. First, this study focused on geometric changes to the CNS-OARs. The interaction between target and OARs and the dosimetric, local control and toxicities from chemoIMRT were not included. Second, the OAR-individualized PRV margin for head and neck cancer sounded reasonable but faced many challenges in the clinical practice. Third, CT images without contrast might be insufficient to clearly delineate the CNS-OARs, especially these complicated structures locate in base of the skull and magnetic resonance imaging might be more accurate. The reported time-dependent geometric changes to the CNS-OARs during IMRT require validation in future studies. Lastly, although rigid registration was used to mimic the execution of imaging-guided IMRT in our daily clinical practice, this imaging registration might unavoidably result in some matching error, which was not included this study. Though these disadvantages, our findings could provide some useful information to refine the radiotherapy of head and neck cancer, especially for NPC.

## 5. Conclusions

The CNS-OARs in patients with NPC may undergo considerable volume and shape changes in the context of chemo-IMRT. The volume of brainstem and spinal cord decreased over time with consecutive RT fractions. And the volume change of specific organs was correlated with patients’ weight loss. For most CNS-OARs, the shape of the CNS-OAR also experienced considerable changes due to several factors. Moreover, the positional displacement of spinal cord in the lower neck section was much larger than tha in the upper neck. These quantifiable geometric changes might be used to improve IMRT and ART protocols for head and neck cancer. Future studies should investigate the impact of these geometric changes on radiation dose distribution, the sparing of RT-related toxicities and the loco-regional control.

## Data Availability

The datasets used and/or analyzed during the current study are available from the corresponding author on reasonable request.

## Acknowledgments

We are grateful to Prof. Jan-Jakob Sonke from the Department of Radiation Oncology, The Netherlands Cancer Institute, Amsterdam, The Netherlands, who generously provided research software (WRLDMATC) developed in-house for this type of research. We would like to thank Editage Inc. for their careful English language editing and proofreading.

